# The impact of the COVID-19 pandemic on mental health care utilization among people living with HIV: A real-world data study

**DOI:** 10.1101/2024.09.26.24314443

**Authors:** Atana Pasha, Shan Qiao, Jiajia Zhang, Ruilie Cai, Buwei He, Xueying Yang, Chen Liang, Sharon Weissman, Xiaoming Li

## Abstract

**Objective:** The COVID-19 pandemic has profoundly impacted mental health worldwide, particularly among vulnerable populations such as people living with HIV (PLWH). However, large-scale, real-world data on mental health care utilization and associated factors among PLWH remain limited. This study leveraged electronic health records (EHR) and Basics survey data from the All of Us program to explore mental health care utilization and associated factors among PLWH during the COVID-19 pandemic.

**Methods:** Using a retrospective cohort design, we identified and included 4,575 PLWH through computational phenotyping based on relevant Observational Medical Outcomes Partnership (OMOP) Common Data Model (CDM) concept sets from the All of Us program between March 2018 and March 2022. Mental health care utilization was measured using the yearly count of mental health care visits over this period. The pattern of mental health care utilization was compared between pre-pandemic (2018-2020) and during the pandemic (2020-2022). Incidence rate ratios (IRR) from the Poisson generalized linear mixed models (GLMM) were used to examine associations between mental health care utilization, history of COVID-19 infection, demographic factors, pre-existing chronic conditions (e.g., hypertension, diabetes), and socioeconomic status.

**Results:** Among 4,575 PLWH, the annual number of mental health care visits decreased significantly during the pandemic period (March 2020 – March 2022) compared to the pre-pandemic period (March 2018 – February 2020) (IRR = 0.89, p < 0.001). The Poisson regression analysis found that a history of COVID-19 infection was associated with a higher number of mental health care visits (IRR = 1.35, p < 0.001). Middle-aged groups with participants aged 30-39 (IRR= 2.35, p = 0.002), 40-49 (IRR= 3.49, p < 0.001), and 50-64 (IRR= 2.07, p = 0.004) had significantly higher visit numbers compared to the youngest group (18-29 years). Black or African American participants were less likely to have visits compared to White participants (IRR = 0.71, p = 0.002). Medicaid health insurance was associated with an increase (IRR = 1.29, p = 0.007), while employer- or union-sponsored insurance was associated with a decrease in mental health care utilization (IRR = 0.54, p < 0.001, p < 0.001). Pre-existing comorbidities greatly increased the number of mental health care visits (one comorbidity: IRR = 5.49, two or more: IRR = 10.4, p < 0.001).

**Conclusion:** Our study reveals a significant decrease in mental health care utilization and the diverse experiences of mental healthcare among PLWH during the COVID-19 pandemic. These findings underscore the importance of addressing disparities in mental health care access, particularly during public health emergencies, and suggest the need for tailored interventions to meet the mental health care needs of PLWH.

**Key Points:** *Question:* How did the COVID-19 pandemic affect mental health care utilization among people living with HIV (PLWH)? What are the factors associated with mental care health utilization among PLWH across both the pre-pandemic and pandemic periods?

*Findings:* Real-world data from a nationwide cohort of 4,575 PLWH identified from the All of Us program in the United States between March 2018 and March 2022 were used to investigate the effect of COVID-19 pandemic on mental health care utilization (i.e., the yearly count of mental healthcare visits). There was a considerable decrease in mental health care utilization during the pandemic (2020-2022) compared to the pre-pandemic period (2018-2020). Factors associated with higher mental health care utilization during the pandemic were the experience of COVID-19 infection, being middle-aged, Medicaid health coverage, and having comorbidities. Black African American participants, those with employer- or union health insurance, and high annual income (>$35,000), had lower mental health care utilization.

*Meaning:* Our study highlights the diverse and unequal experiences of PLWH during the pandemic, with variations in mental health care utilization over time and across different disadvantaged subgroups. These findings underscore the need for timely and targeted mental health care services and delivery in future public health crises.

## INTRODUCTION

The COVID-19 pandemic has not only resulted in high mortalities and morbidities but also profoundly affected people’s mental health, especially for vulnerable populations such as people living with HIV (PLWH) (1–3). Psychiatric disorders, such as depression, psychosis, substance use disorder, and post-traumatic stress disorder, are 1.5 to 8 times more common among PLWH than in the general population (4). The prevalence of depressive disorder is approximately 1.5 to 2.5 times higher in PLWH compared to the general population (5, 6). Studies have also shown higher levels of anxiety and suicidality among PLWH (7–9). The increasing mental health challenges faced by PLWH stem from the existing physical health burdens of HIV, the heightened risk of severe illness from COVID-19, and other structural-level adversities and vulnerabilities. For instance, HIV-related services disruption, fear of comorbidities with COVID-19, the dual burdens of these diseases, social isolation, limited access to healthcare services, increased stigma, and financial problems result in poor mental health among PLWH (1, 10–16).

Therefore, the prioritization of addressing mental health among PLWH during the COVID-19 pandemic has been identified as a crucial aspect of the response to this public health emergency by the Office of AIDS Research (OAR) and the National Institute of Mental Health (NIMH) (17). Unfortunately, the difficulties in receiving care were especially evident during the pandemic, as many PLWH reported difficulties in accessing and using mental health care, including in scheduling, appointment availability, and the use of telemedicine, which may not have been available to all patients because of the digital divide (13–16). The under-optimized utilization of mental health care services can lead to worsened psychosocial well-being and HIV-related outcomes, such as increased anxiety, depression, and decreased adherence to HIV treatment and follow-up (13, 18–20). The negative impacts of COVID-19 pandemic on mental health care utilization among PLWH are most significant among minority and underserved populations who already experience disparities in healthcare access (13, 21–27).

Existing literature has demonstrated how the COVID-19 pandemic has further aggregated the preexisting mental health disparities. For example, recent studies report increased alcohol consumption, binge drinking, and recreational drug use during the pandemic among men who have sex with men (MSM), including those living with HIV (28). However, the number of empirical studies focusing on mental health care service interruption and its disparities during PLWH was small, and most studies were usually conducted within an existing HIV-related intervention cohort or cross-sectional surveys. These studies were limited by a small sample size and biases due to self-reported measures. ICD-10 codes in electronic health records (EHR) provide psychiatric disorder diagnoses (mood disorders, anxiety disorders, substance use disorders, stress-related disorders, schizophrenia, and personality disorders), which can be a proxy measure of mental health care visits. The large cohort of PLWH and rich data in the NIH’s Cloud-based All of Us program have presented a unique opportunity to address these research gaps. Therefore, this study used real-world data from a large, diverse cohort of PLWH from the All of Us program to address the following research questions:

- How did mental health care utilization change before and during the COVID-19 pandemic?
- What factors are associated with mental health care utilization among PLWH across both the pre-pandemic and pandemic periods?

Examining the disruptions of the COVID-19 pandemic and associated factors will help assess the extent of mental health care challenges experienced by PLWH during the pandemic, identify the most vulnerable sub-populations enduring mental health disparities, and inform strategies on how to improve continuity of care and mental health support during future public health emergencies.

## METHODS

### Study Design and Data Source

This study used a retrospective cohort design to examine the impact of the COVID-19 pandemic on mental health care utilization among PLWH in the United States. The All of Us program aims to enroll over 1 million diverse participants aged 18 and older in the US and has collected various types of data since 2018. The electronic health records (EHR) data used in the study were longitudinal data extracted from 2018 to 2022 and were a key source of outcome variable (yearly count of mental health care visits). The study cohort included PLWH identified through computational phenotyping based on the EHR data and Personal and Family Health History data. The deidentified data are available to researchers through public, controlled, and registered tiers. For the Registered Tier that we used in this study, EHR data are normalized by the Observational Medical Outcomes Partnership (OMOP) Common Data Model (CDM). Additional personal data collected from Survey data consisted of the Basics survey (28 questions), which asks for information on basic demographics, household size, health insurance, disability status, employment and income, and housing conditions. The Overall Health survey (21 questions) focuses on health literacy, general health conditions, women’s health issues (if applicable), and transplant history. Additional surveys on specific subjects (e.g., personal medical history, family medical history, and health access and utilization) are added regularly. Data from these surveys were used as baseline information and have been normalized by OMOP.

### Study Cohort

The study cohort included PLWH identified through computational phenotyping based on the EHR data and Personal and Family Health History data (29). The individuals in the study cohort could be enrolled in the All of Us database at any time from May 2017 to March 2022. We modified the concept sets given the All of Us project contains survey data in addition to EHR data. The logical procedures for phenotyping the PLWH cohort were followed by the longitudinal study protocol Using All of Us data to examine the mental health change during the COVID-19 pandemic among PLWH (17).

### Outcome Measures

In this paper, we measured mental health care utilization using the yearly counts of mental health care visits in the All of Us Research Program, based on ICD-10 codes for mental health disorders across 6 broad domains: mood disorders, anxiety disorders, substance use disorders, stress-related disorders, schizophrenia, and personality disorders (30). The number of mental health related visits was divided into four intervals, which tracks the number of mental health care visits across two 2-year intervals (March 2018 -February 2020 as pre-pandemic period and March 2020 - March 2022 as pandemic period). We analyzed the overall trend in mental health care utilization over these intervals and assessed factors associated with mental health care utilization during the pandemic.

### Covariates

A wide range of demographic, socioeconomic, and health-related factors were considered as covariates in the analysis to understand their association with mental health care utilization among PLWH during the COVID-19 pandemic. Demographic variables included age, gender, race/ethnicity, marital status, and sexual orientation. Socioeconomic variables comprised household income, educational attainment, and type of health insurance, such as Medicaid or employer-sponsored insurance. Health-related covariates involved the history of COVID-19 infection, the number of pre-existing chronic conditions such as hypertension and diabetes, and HIV-related health indicators.

### Statistical Analysis

We employed Poisson generalized linear mixed models (GLMM) to assess the yearly count of mental health care visits. Incidence rate ratios (IRR) were calculated to evaluate the relative change in the number of mental health visits across different intervals and among various demographic, socioeconomic, and health-related subgroups. The IRR quantified and demonstrated differences in the number of mental health visits in four distinct intervals, accounting for the effects of covariates. An IRR greater than 1 indicates an increased number of visits, while an IRR less than 1 indicates fewer visits.

## RESULTS

### Characteristics of participants

We identified a cohort of 4,575 PLWH, while 3042 (66%) were males, 1348 (29%) were females, and about 185 (4%) were identified as not man only, not women only, preferred not to answer, or skipped the gender question (Table 1). The average age of the cohort was 55 (SD=12.44 years). Half of the participants were in the age group of 50-64 years (50%), followed by the age group of 65+ years (20%), 40-49 years (15%), 30-39 years (12%), and 18-29 years (3%). Black/African Americans were the largest racial group in the study (49%), followed by Asian/others/unknown (28%) and White (22%). In terms of ethnicity, nearly 18% of the cohort were Hispanic or Latino. About 46% of the cohort were never married, and 45% were identified as straight in terms of sexual orientation. Regarding socioeconomic status, 91% of the PLWH had a high school degree or more, and 59% had an annual income of less than $35,000. Regarding health insurance, 48% had Medicaid, 24% had Medicare, 13% had employer- or union-sponsored insurance, and 11% claimed other health insurance plans. With aspects of pre-existing health conditions, nearly 13% of the cohort reported disabilities, 20% of them reported one chronic disease besides HIV, and 18% reported two or more types of comorbidities. Nearly 11% of the cohort reported a history of COVID-19 infection.

**Table 1.**
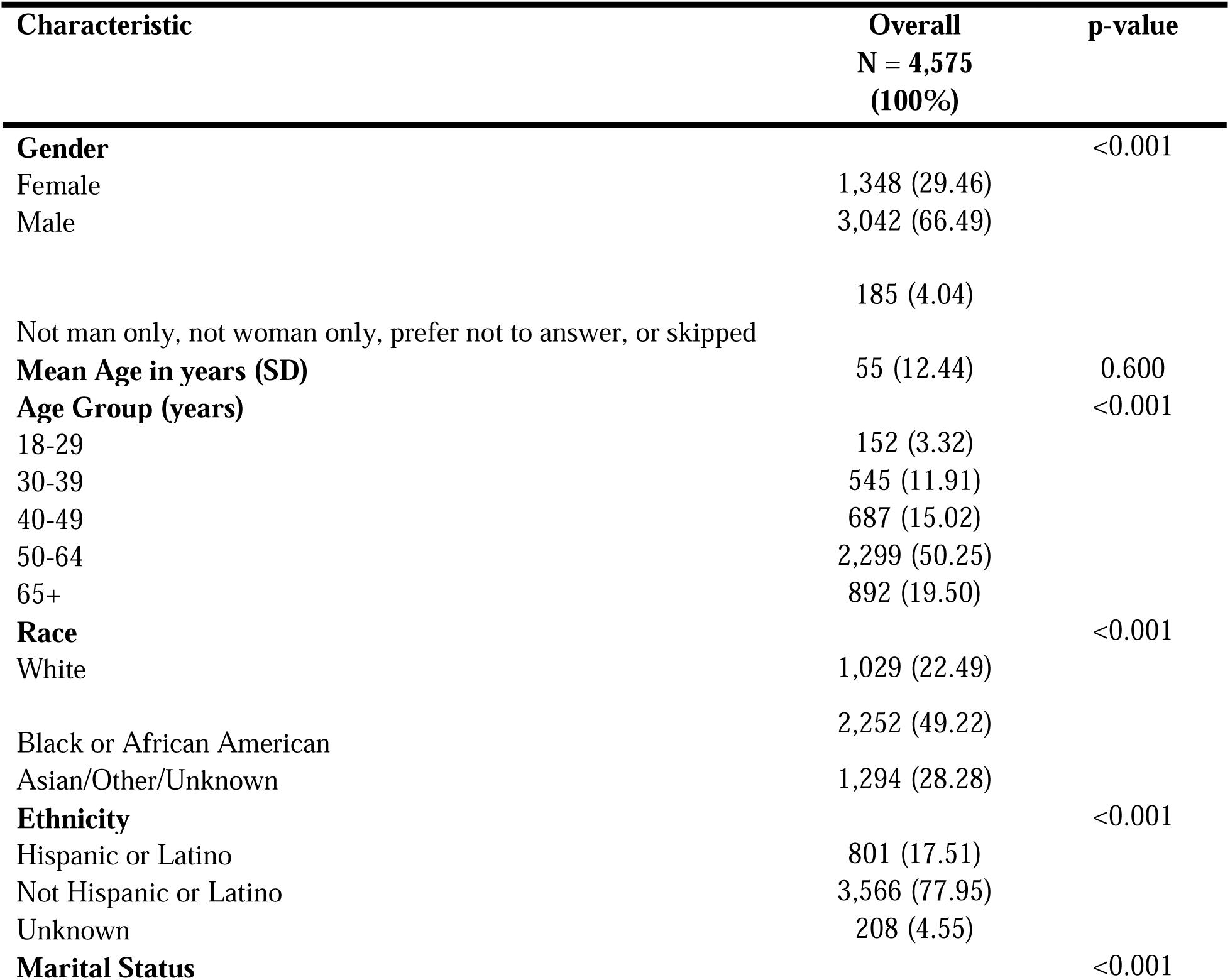
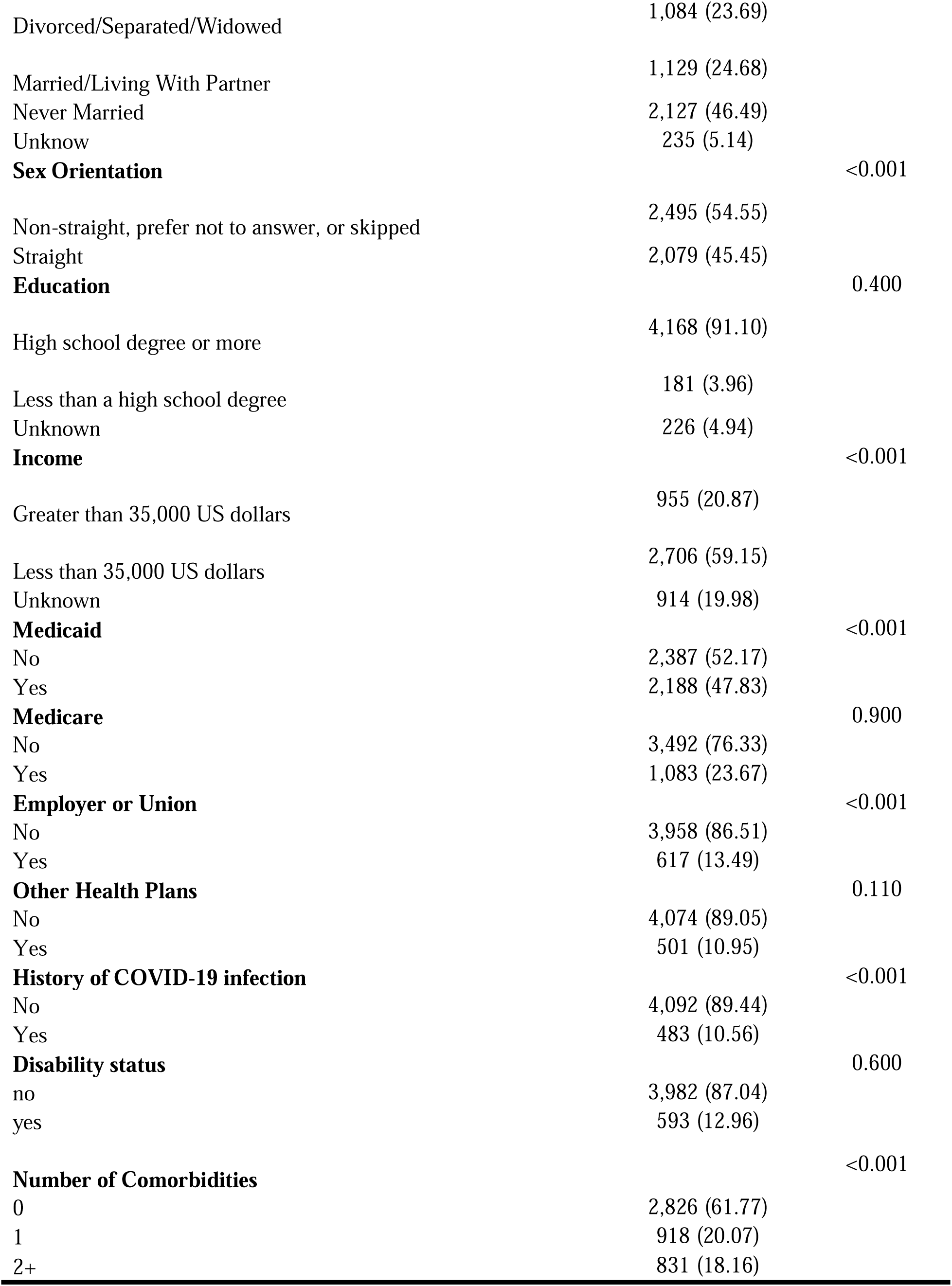
Summary of Demographic Characteristics of the People Living With HIV (PLWH) Cohort.

### Poisson GLMM Analysis

The yearly count of mental health care visits, assessed using Poisson GLMM, showed a significant decrease during the pandemic (March 2020 – March 2022) compared to the pre-pandemic (March 2018 – February 2020) (IRR = 0.89, p < 0.001). Our study also found several factors that were significantly associated with mental health care utilization among PLWH during the COVID-19 pandemic (Table 2).

**Table 2.** Poisson Generalized Linear Mixed Models (GLMM) of Factors Associated with Mental Health care utilization, Including History of COVID-19 infection, Age, Income, Insurance, and Number of Comorbidities.

| Characteristic | IRR1 | 95% CI1 | p-value |
| --- | --- | --- | --- |
| **History of COVID-19 Infection** |  |  |  |
| Status: No | — | — |  |
| Status: Yes | 1.35 | 1.28, 1.42 | <0.001 |
| **COVID outbreak** |  |  |  |
| 0 | — | — |  |
| 1 | 0.89 | 0.87, 0.90 | <0.001 |
| **Gender** |  |  |  |
| Female | — | — |  |
| Male | 1.04 | 0.84, 1.28 | 0.7 |
| Not man only, not woman only, prefer not to answer, or skipped | 1 | 0.63, 1.60 | >0.9 |
| **Age group** |  |  |  |
| 18-29 | — | — |  |
| 30-39 | 2.35 | 1.38, 4.02 | 0.002 |
| 40-49 | 3.49 | 2.07, 5.88 | <0.001 |
| 50-64 | 2.07 | 1.25, 3.41 | 0.004 |
| 65+ | 1.24 | 0.72, 2.12 | 0.4 |
| **Race group** |  |  |  |
| White | — | — |  |
| Asian/Other/Unknown | 0.81 | 0.54, 1.19 | 0.3 |
| Black or African American | 0.71 | 0.57, 0.89 | 0.002 |
| **Ethnicity group** |  |  |  |
| Not Hispanic or Latino | — | — |  |
| Hispanic or Latino | 1.32 | 0.90, 1.94 | 0.2 |
| Unknown | 1.13 | 0.65, 1.96 | 0.7 |
| **Marital Status** |  |  |  |
| Never Married | — | — |  |
| Divorced/Separated/Widowed | 1.21 | 0.98, 1.49 | 0.081 |
| Married/Living with Partner | 0.87 | 0.71, 1.08 | 0.2 |
| Unknown | 1.26 | 0.85, 1.86 | 0.3 |
| **Sexual Orientation** |  |  |  |
| Straight | — | — |  |
| Non-straight orientation, prefer not to answer, or skipped | 1 | 0.82, 1.22 | >0.9 |
| **Education** |  |  |  |
| Less than a high school degree | — | — |  |
| High school degree or more | 1.25 | 0.83, 1.90 | 0.3 |
| Unknown | 1.17 | 0.67, 2.06 | 0.6 |
| **Income** |  |  |  |
| less than 35,000 US dollars | — | — |  |
| greater than 35,000 US dollars | 0.47 | 0.36, 0.61 | <0.001 |
| Unknown | 0.7 | 0.56, 0.87 | 0.001 |
| **Disability** |  |  |  |
| No | — | — |  |
| Yes | 1.02 | 0.80, 1.31 | 0.9 |
| **Medicaid** |  |  |  |
| No | — | — |  |
| Yes | 1.29 | 1.07, 1.55 | 0.007 |
| **Medicare** |  |  |  |
| No | — | — |  |
| Yes | 0.8 | 0.65, 0.99 | 0.042 |
| **Employer or Union** |  |  |  |
| No | — | — |  |
| Yes | 0.54 | 0.39, 0.74 | <0.001 |
| **Other Health Plans** |  |  |  |
| No | — | — |  |
| Yes | 1.17 | 0.88, 1.55 | 0.3 |
| **Number of Comorbidities** |  |  |  |
| 0 | — | — |  |
| 1 | 5.49 | 4.46, 6.77 | <0.001 |
| 2 and more | 10.4 | 8.30, 12.9 | <0.001 |

Factors associated with higher mental health care utilization among PLWH included a history of COVID-19 infection, with participants who had COVID-19 infection showing a higher number of mental health care visits (IRR = 1.35, p < 0.001). Older age was another key factor, with participants aged 30-39 (IRR= 2.35, p = 0.002), 40-49 (IRR= 3.49, p < 0.001), and 50-64 (IRR= 2.07, p = 0.004) having a significantly higher number of mental health care visits compared to the youngest group (18-29 years). People with Medicaid health insurance had a higher number of mental health care visits (IRR= 1.29, p = 0.007). Pre-existing comorbidities associated with an increased number of visits, with one comorbidity (IRR = 5.49, p < 0.001) or two or more comorbidities (IRR = 10.4, p < 0.001) significantly elevating the yearly count of mental health care visits.

On the other hand, several factors were associated with a decrease in the yearly count of mental health care visits. High income was associated with a decrease in a yearly count of mental health care visits, as participants with an annual income higher than $35,000 had fewer mental health care visits (IRR = 0.47, p <0.001). Race played a role, with Black or African American participants (IRR = 0.71, p = 0.002) less likely to have mental health care utilization compared to White participants. Health insurance type was another important factor, as participants with employer- or union-sponsored health insurance had fewer mental health care visits (IRR = 0.54, p < 0.001).

## DISCUSSION

The present study demonstrated the significant impact of the COVID-19 pandemic on mental health care utilization among PLWH. The reduction in mental health care utilization during the pandemic, which was seen in our study, is in line with previous studies that have documented limited access to care and reduced healthcare seeking behavior among PLWH during the COVID-19 pandemic (1, 10, 19, 31–33). According to the World Health Organization (WHO), essential mental health services were interrupted in more than 90% of the countries during the time of social isolation (34). When social distancing measures were implemented, and the health care systems were under pressure, the availability of basic mental health services was reduced, especially for vulnerable groups such as the PLWH (11). This disruption of the healthcare services resulted in reduced access to mental health services (19). Some of the PLWH have avoided in-person follow-up appointments because of COVID-19 related concerns (11, 20, 35–38). Telemedicine became essential during the pandemic, acting as a way of ensuring that many PLWH could continue to access mental health services (14, 39). However, challenges, including the digital divide, inadequate access to technology, and issues of privacy, limited the access to telemedicine for some PLWH, especially the older PLWH and the economically disadvantaged people (1, 10, 14, 15, 40).

The number of mental health care visits among PLWH on an annual basis during the COVID-19 pandemic was associated with several factors. History of COVID-19 infection, middle age, Medicaid health coverage, and comorbidity (higher in multiple comorbidity) were associated with an increase in the yearly count of mental health care visits among PLWH during the pandemic. Participants who self-identified as Black or African American, those with employer- or union-sponsored insurance, and high income had fewer mental health care visits. These findings are consistent with previous studies that indicated that economic vulnerability and comorbid conditions increase the burden of mental health among PLWH (1, 20, 32, 41, 42). Financial difficulties combined with HIV-related prejudice made some people unable to receive treatment and, consequently, fail to attend their appointments (19).

We identified pre-existing comorbidities, particularly multiple comorbidities, associated with an increase in mental health care utilization among PLWH, which is consistent with previous studies (43). The presence of multiple comorbidities increased the mental health burden on PLWH during the pandemic, as managing various chronic conditions in the context of a global health crisis proved to be a significant source of stress (18). Coping with various stressful situations might have adverse consequences for mental health (44). In this regard, the history of COVID-19 infection adds a layer of health-related anxiety and potential stigma, and the stress of managing two or more chronic conditions, social isolation during the COVID-19 pandemic, or economic and societal disruptions caused by the pandemic may contribute to the increase in the yearly counts of mental health care visits (13, 45).

Our study results also indicate that aging with HIV is associated with an increase in mental health care utilization during the COVID-19 pandemic. Older PLWH, in particular, faced significant challenges during the pandemic, as physical distancing measures worsened their feelings of isolation and loneliness, contributing to poorer mental health outcomes (13, 43, 46). Previous research indicates that older adults and those with underlying health conditions are at greater risk for severe illness from COVID-19, which may have heightened their anxiety and reluctance to seek in-person care (2, 13, 43).

On the other hand, some factors were linked with a decrease in mental health care utilization among PLWH during the COVID-19 pandemic. Employer- or union-sponsored insurance and Black or African American participants were less likely to have had a mental health care visit than other participants, which could be due to access issues or the ways in which care was sought during the pandemic. It has been established that cultural mistrust and perceived discrimination are potential barriers to mental health care service use among Black or African Americans (1, 11, 21, 27). The COVID-19 pandemic has also highlighted the importance of health insurance in providing adequate health care. In our study, having Medicaid as one of the health insurance options was an associated factor for a higher number of mental health care visits, while having employer- or union-sponsored health insurance was associated with a decrease in mental health care utilization. Employer- and union-sponsored health insurance is a sign of job security, which can be considered as a buffer against other living and care expenses stress (1, 19, 32). On the other hand, Medicare health coverage may be related to low-income levels. The ability to afford care and the availability of preventive health visits play pivotal roles in managing HIV as a chronic infection, as health insurance coverage disparities may affect an individual’s likelihood to seek healthcare and impact HIV outcomes (9, 47–50).

Several strengths can be identified in our study. Real-world data from a large diverse cohort of PLWH from the All of Us program (EHR and survey data) were used to improve the validity of the findings on mental health care utilization among PLWH during the COVID-19 pandemic. The use of pre-pandemic data (2018–2020) enabled us to compare changes in the yearly counts of mental health care visits before and during the COVID-19 pandemic. Furthermore, we identified factors that are associated with the yearly count of mental health care visits among PLWH, contributing to tailoring interventions for the most vulnerable subpopulations facing mental health disparities. However, the present study has some limitations, including some characteristics used survey data that depended on self-reports and were therefore susceptible to recall and social desirability bias. Additionally, the unavailability of data on telehealth services for mental health care during the pandemic presents a limitation, as telehealth became a prominent mode of care delivery during this period. Finally, although data before the pandemic was incorporated, more follow-up data are required to determine the long-term trend of mental health care utilization among PLWH.

## CONCLUSION

Our study revealed a significant decrease in mental health care utilization among PLWH during the COVID-19 pandemic, highlighting the interruption of mental health care services. Factors such as being middle-aged, experiencing COVID-19 infection, Medicaid coverage, and pre-existing comorbidities were associated with an increased number of yearly mental health care visits. In contrast, Black or African American participants and those having employer- or union-sponsored health insurance and high income had fewer mental health care utilization. These results underscore the importance of developing targeted strategies to maintain PLWH’s access to mental health care, especially during a public health crisis among vulnerable sub-populations. It is crucial to embrace a comprehensive and multisectoral approach to mental health and HIV care. Leveraging mHealth and other technology-based interventions can be efficient when in-person access is not possible. Policies need to consider the socioeconomic and psychological effects of pandemics and make sure that mental health care services are prepared to deal with future public health crises.

## Acknowledgments

The authors thank the All of Us Program.

## Data Availability

The data used in this study are derived from the All of Us Research Program, which is accessible to approved researchers through the All of Us Researcher Workbench. Access to the data requires registration, completion of the appropriate training, and approval of a research project by the All of Us Data and Research Center. For more information on accessing the data, please visit the All of Us Researcher Workbench (https://workbench.researchallofus.org/).

## Declarations

### Ethical Approval

The study has been approved by the institutional review board at the University of South Carolina (Pro00124044).

